# Do the Health Benefits of Boiling Drinking Water Outweigh the Negative Impacts of Increased Indoor Air Pollution Exposure?

**DOI:** 10.1101/2024.03.22.24304348

**Authors:** Emily Floess, Ayse Ercumen, Angela Harris, Andrew P. Grieshop

## Abstract

**Background:** Billions of the world’s poorest households are faced with the lack of access to both safe drinking water and clean cooking. One solution to microbiologically contaminated water is boiling, often promoted without acknowledging the additional risks incurred from indoor air degradation from using solid fuels.

**Objectives:** This modeling study explores the tradeoff of increased air pollution from boiling drinking water under multiple contamination and fuel use scenarios typical of low-income settings.

**Methods:** We calculated the total change in disability-adjusted life years (DALYs) from indoor air pollution (IAP) and diarrhea from fecal contamination of drinking water for scenarios of different source water quality, boiling effectiveness, and stove type. We used Uganda and Vietnam, two countries with a high prevalence of water boiling and solid fuel use, as case studies.

**Results:** Boiling drinking water reduced the diarrhea disease burden by a mean of 1110 DALYs and 368 DALYs per 10,000 people for those under and over <5 years of age in Uganda, respectively, for high-risk water quality and the most efficient (lab-level) boiling scenario, with smaller reductions for less contaminated water and ineffective boiling. Similar results were found in Vietnam, though with fewer avoided DALYs in children under 5 due to different demographics. In both countries, for households with high baseline IAP from existing solid fuel use, adding water boiling to cooking on a given stove was associated with a limited increase in IAP DALYs due to the log-linear dose-response curves. Boiling, even at low effectiveness, was associated with *net* DALY reductions for medium- and high-risk water, even with unclean stoves/fuels. Use of clean stoves coupled with effective boiling significantly reduced total DALYs.

**Discussion:** Boiling water generally resulted in net decreases in DALYs. Future efforts should empirically measure health outcomes from IAP vs. diarrhea associated with boiling drinking water using field studies with different boiling methods and stove types.

## Introduction

Globally, many households face the challenges of both poor drinking water quality and the lack of access to clean cooking fuels. Two billion people lack safely managed drinking water.^1^ In 2019, 1.53 million deaths were attributed to diarrheal diseases,^2^ and 60% of all diarrheal deaths are due to improper water, sanitation, and hygiene (WASH).^3^ Diarrhea is the leading cause of death and illness globally among children under 5 years.^4^ Meanwhile, 2.6 billion people cook using solid fuels^5^ resulting in high exposures to fine particulate matter (PM_2.5_).^6^ Indoor air pollution (IAP) causes stroke, ischemic heart disease, chronic obstructive pulmonary disease, lung cancer,^7^ and acute lower respiratory infections.^8^ IAP was associated with 6.7 million premature deaths annually in 2019.^9^

A third of households in 67 countries report treating their drinking water at home.^10^ Boiling water is the most common household water treatment,^11^ with an estimated 1.2 billion users (70% of all household water treatment users).^12^ Boiling for water treatment is most common in the Western Pacific region and least common in the Eastern Mediterranean and African regions. It is widespread in many Asian nations including Indonesia (90.6% of households practicing water treatment methods reporting boiling), Mongolia (95.2%), Uzbekistan (98.5%) and Vietnam (91.0%).^11^ Though boiling on the African continent is comparatively less common, many countries in Africa have high rates of boiling,^11,13^ including Lesotho, Rwanda, Uganda (more than 80% of households reported boiling), and Burundi and Namibia (over 60% reported boiling).^13^ Boiling for water treatment has been widely promoted for decades for low-income countries and emergency situations.^14^ Limitations of boiling include potential recontamination of stored boiled water by contact with hands and utensils because boiling does not provide residual protection. Improper boiling methods can also result in poor water quality^15–17^ and families often mix boiled and non-boiled water.^18^

Other concerns raised by boiling water are the potential for increased air pollution exposures from fuel combustion^19^ and high fuel costs.^20^ Solid fuel use is prevalent in low-income settings for both cooking and boiling water. Reducing the use of solid fuels reduces indoor fine particulate matter (PM_2.5_) concentrations,^21^ yet transitions to clean fuels (e.g., liquified petroleum gas; LPG) have proven challenging.^22^ Despite the perception that clean fuels are better and more convenient,^23^ the associated financial burden (e.g., the cost relative to wood, which is often cheap or free) and other barriers often prevent their widespread adoption.^24^ One proposed solution is “improved cooking solutions” (ICS), or “*Cooking solutions that improve, however minimally, the adverse health, environmental, or economic outcomes from traditional solid fuel technologies”.*^25^ ICS includes natural- or forced-draft biomass cookstoves with improved combustion efficiency. For example, pellet-fed gasifier stoves emit >90% less (PM_2.5_) than conventional stoves.^26–28^ However, despite their potential to provide health and other benefits relative to traditional stoves, behavioral, technical, and economic challenges have limited their adoption.^29,30^ Electric stoves are an additional clean option but are not common in low-income countries due to their cost and the poor availability and reliability of electrical grid connections. In Uganda, for example, less than 1% of the population cooks with electricity^31^ due to the expense of electricity and lack of subsidies.^32^

Those working to mitigate risks from indoor air pollution (IAP) and unsafe drinking water face similar challenges in designing, implementing, and securing the sustained use of interventions such as clean fuel and household water treatment.^30^ However, few studies have examined these linked risks together. A randomized controlled trial in Rwanda combining a cookstove and water filter intervention found the intervention reduced the prevalence of reported child diarrhea by 29% and that the benefits of the program outweighed the financial costs.^33,34^ In a related cost-benefit analysis, the averted DALYs from using a water filter and improved cookstove were found to be 239 and 556 per 10,000 people per year, respectively.^33^ However, the study observed no significant reduction in 48-hour personal exposure to PM_2.5_,^34^ consistent with challenges faced by stove replacement programs observed elsewhere.^35^ A study in China measured the reduction in thermotolerant coliforms in water from boiling using different methods and modeled air pollution from boiling water. The modeled mean 24-hour PM_2.5_ kitchen concentration from boiling water with biomass combustion was 79 µg/m^3^ ^19^, substantially above the WHO interm-1 target of 35 µg/m^3^.^36^ Boiling with electric kettles was associated with the largest reduction in thermotolerant coliforms. However, the study did not measure health outcomes. To date, no study has specifically investigated the tradeoffs associated with drinking water treatment by boiling using solid fuels and compared the health risks. The overarching goal of this study is to develop a modeling framework to quantify the net health impacts from boiling drinking water with solid fuels, accounting for a range of IAP-associated health outcomes and for diarrhea associated with water contaminated with fecal matter. This framework is then applied using available literature value for inputs for two countries, Uganda and Vietnam, selected as case studies.

## Methods

### Framework Definition and Test Population

DALYs are commonly used to quantify health burdens because they account for morbidity with differential disease severity^37^ and mortality. In our study, we used DALYs as the primary metric to compare multiple risks.^38^ Quantitative Microbial Risk Assessment (QMRA) models are commonly used to determine the diarrhea risk associated with consuming water from a particular water source.^39^ For IAP, the population attributable fractions based on dose-response curves for individual diseases are used to calculate the burden of disease.^40,41^

We adopted these two methods (Figure 1), creating two modules, and used literature-derived distributions of the relevant IAP, QMRA, and demographic parameters as inputs (See Supplemental Material, Parameters Needed for Model). The *water risk module* uses a QMRA model to calculate the DALYs from drinking water contaminated by fecal matter before and after treatment by boiling. The *air risk module* uses an indoor box model to quantify the PM_2.5_ concentrations for different stoves and the Household Air Pollution Intervention Tool (HAPIT)^41^ to quantify the DALYs associated with IAP under various scenarios. Both modules employ Monte Carlo simulations to capture the influence of variability and uncertainty in input parameters on model outputs.

**Figure I.**
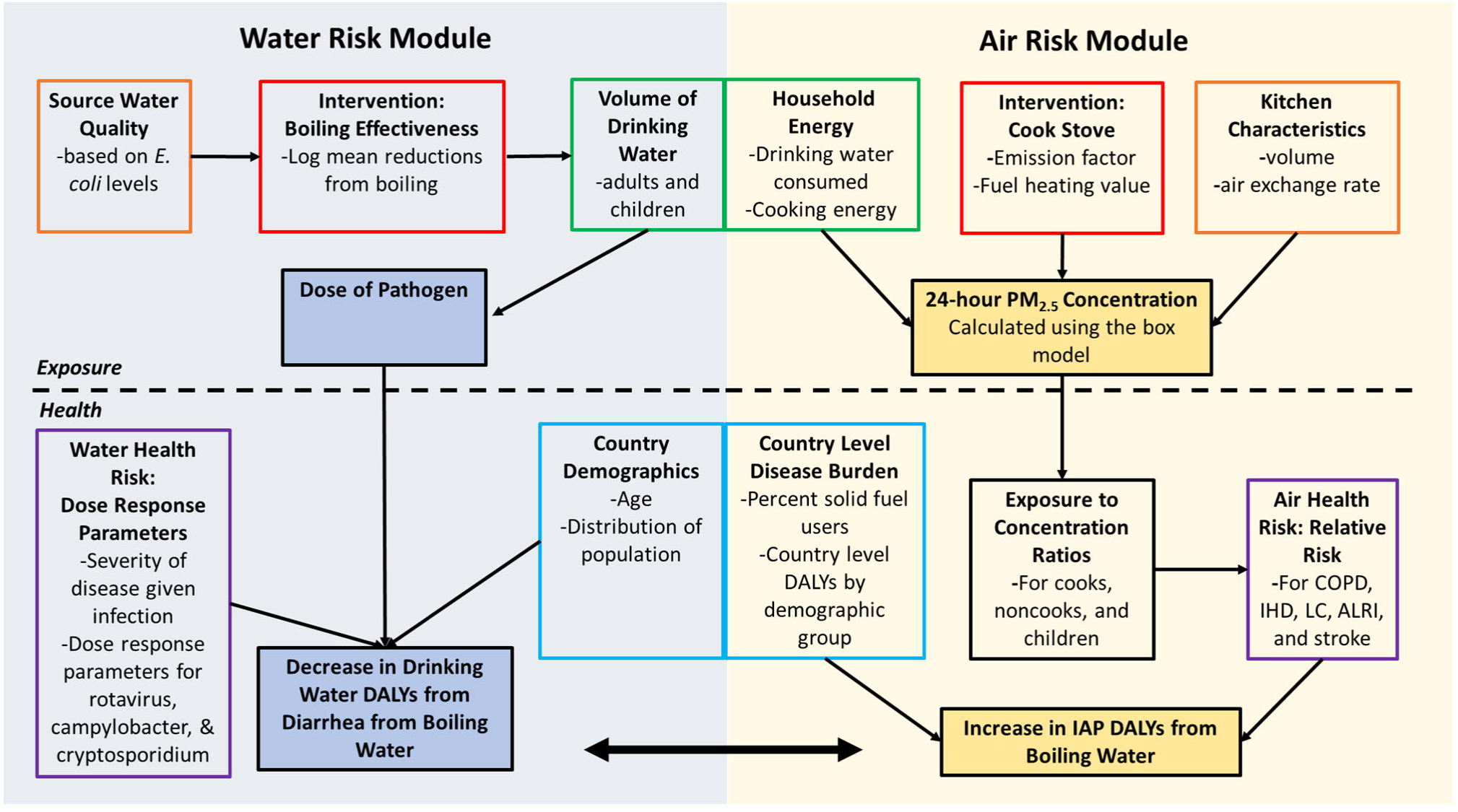
Overview of methods to calculate and compare drinking water DALYs and /AP DALYs

Globally, more than half of all deaths from IAP from solid fuel are from acute lower respiratory infection (ALRI) in children under 5 years of age^42^ and most diarrheal DALYs occur in children under 5 years of age.^43^ Therefore, we conducted analyses for two age groups: those under 5 years of age (“under-5s”), and all other age groups combined (“over-5s”).

We designed the model to be used for any setting. However, we selected Uganda and Vietnam as case study countries as they are in distinct regions, have different population demographics, and high prevalences of boiling among household water treatment users (82% in Uganda, 91% in Vietnam) and solid fuel use (96% in Uganda, 35% in Vietnam).^5,11,13^ We assumed a household size of 5 people per household for both countries and that each household had one cook.^41,44^ The number of children under 5 per household varied between the two countries (1 in Uganda, 0.4 in Vietnam), based on the values used in the HAPIT model (See Supplemental Material, Household Demographics for Vietnam and Uganda).^41,44^ The models were run for a sample population of 10,000 people.

### Health Benefits from Boiling Drinking Water

The risk of illness from contaminated drinking water is characterized using a QMRA, focusing on selected reference pathogens.^45^ Pathogens used in QMRAs are typically selected based on global public health relevance; because they are transmitted via environmental, waterborne, and foodborne routes,^46^ the dominant pathogens may also depend on geographic region.^47^ For this study, we selected pathogens identified as leading causes of diarrhea in multisite studies of diarrhea etiology in low-income countries.^48–50^ Based on these studies, the locations selected for our modeling exercise, and previous QMRA studies,^51–53^ we selected a virus (rotavirus), a protozoan (*Cryptosporidium*) and a bacterium (*Campylobacter*) to quantify the risk from exposure to fecally contaminated water.

We used a uniform distribution of *E. coli* levels for each water quality category, using most probable number (MPN) ranges of: 0 MPN/100 mL for ‘safe’ water, 1-10 MPN/100 mL for ‘low-risk’ water, 11-100 MPN/100 mL for ‘medium-risk’ water, and 101-1000 MPN/100 mL for ‘high-risk’ water.^54^ These untreated drinking water categories define baseline water qualities for the model. We used fecal indicator bacteria to pathogen ratios for the three pathogens (See Supplemental Material, Ratio of *E. coli* to Campylobacter, Cryptosporidium, and Rotavirus) from the literature to estimate the abundance of the selected pathogens in untreated water.

We then estimated boiling effectiveness for the selected pathogens. The modeled intervention was boiling at different levels of microbiological effectiveness to account for different field conditions and household practices. The boiling effectiveness was quantified using log-reduction values (LRV), defined as the base-10 logarithm of the ratio of influent to effluent pathogen concentrations. The effectiveness of boiling drinking water was compiled from a literature review of field and lab studies; See Supplemental Material, Log Reduction Values of *E. coli*, TTC, and FC. Water boiling studies reported LRVs for different fecal indicator organisms, including *E. coli,* thermotolerant coliforms (TTC), and fecal coliforms (FC). Since our modeled water quality was characterized using *E. coli,* we converted TTC and FC LRVs to *E. coli* (EC) LRVs^55^ based on assumed ratios. Although *E. coli* to FC ratios can vary in different seasons,^56^ the ratios were assumed to be constant throughout the year. LRV values in the literature (converted to *E. coli*) ranged from 6 for lab-level boiling to -0.26 for ineffective boiling. We multiplied *E. coli* LRVs by a pathogen-specific factor to quantify removal of the respective pathogens (C*ryptosporidium*, C*ampylobacter*, and rotavirus) (See Supplemental Material, Effect of Boiling on *E. coli*, Campylobacter, Cryptosporidium, and Rotavirus).

We used Equation 1 to calculate the daily exposure to pathogens (E_d,_ in MPN),^57^

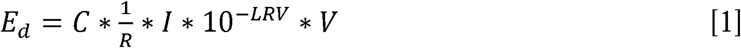

where C is the concentration of pathogen in the source water (MPN per liter), R is the fractional mean analytical recovery (to account for success in microorganism counts) of the pathogen in the sample, I is the fraction of pathogens that are infectious,^51,57^ LRV is the pathogen-specific log-reduction by boiling water, and V (liters) is the daily volume of water consumed per person. We assumed I and R to be 1.

We calculated the probability of illness from the daily exposure using dose-response relationships from the literature.^51,52,58–60^ The dose-response parameters were assumed to be the same regardless of age (See Supplemental Material, Dose-Response Parameters for the Selected Pathogens).

We calculated the probability of developing disease (diarrhea) per single exposure (P_D_) as the product of the probability of disease given infection (P_DI_) and the probability of infection (P_i_) (See Supplemental Material, Probability of Illness Given Infection and Percent Susceptible). The probability of developing illness over period n, (P_D,n_), was calculated using the probability of disease (P_D,n_=1-(1-P_D_)^n^) and the annual symptomatic cases (Cases_year_) was calculated by multiplying the yearly probability of developing illness (P_D,yearly_) times the exposed population.^53,57^

DALYs from diarrhea (years lived with disability) were calculated by taking the disability weight times the duration of disease times the probability of disease.^52,53,57^ Disability weights to quantify disease severity were obtained from the literature (See Supplemental Material, Burden of Disease for Each Pathogen). When calculating years of life lost, the disability weight is 1 and the duration was the life expectancy minus the age at death. The remaining life expectancy at the time of death was randomly selected based on the population age distribution in that country, separated into two categories, children <5 years and all other ages. For disease burden, severity and duration varied by pathogen (*Cryptosporidium*, *Campylobacter*, or rotavirus).^52^

### Health Impacts from Indoor Air Pollution

In the air risk module, first an air pollution box model was used to estimate the 24 hour PM_2.5_ concentrations in the household^61^ and then DALYs were estimated using the HAPIT model.^41^ For the air risk module, we consider two baseline scenarios: one with a household ‘already cooking’ on a traditional woodstove, and one in which a household is ‘not cooking’. We consider the following cookstoves as alternatives against this baseline: 1) improved wood, 2) charcoal, 3) LPG, and 4) electric.^23,62,63^ Electric stoves were included to serve as an ideal counterfactual (i.e., completely clean cooking technology). We considered three categories of stove use: ‘cooking only’, ‘water boiling only’, and ‘cooking and water boiling’.

Our model assumes stove energy is only used for cooking and water heating. The required cooking energy (delivered to pot) for both the Ugandan and Vietnamese households was assumed to be lognormally distributed, with a mean (standard deviation) of 11 (5.5) MJ per day.^61,64^ The daily energy for water heating (E_WH_), (Equation 2), is calculated as the energy needed to heat the water from ambient temperature (T_a_, assumed to be 15 °C) to boiling (100 °C), and then boiled for one minute. The volume of water heated (V, liters) is assumed to be normally distributed with mean (standard deviation) 3.12 (1.17) liters.^65^ This is converted to converted to mass using density (ρ, 1000 g/L). C_p_ is the heat capacity of water (4.186 J g^-1^ K^-1^). Finally, the stove is assumed to boil the water for one minute, so the product of 60 seconds (t) and the power of the stove (P, in Watts) is added to get the total energy demand for heating water in a household.

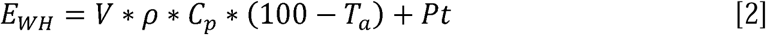

Laboratory studies of boiling effectiveness have been conducted for numerous different temperatures and durations of boiling. Generally, heating water at higher temperatures for longer periods of time results in greater LRVs compared to lower temperatures and shorter periods.^66^ The recommended boiling time in the literature ranges from 1 to 25 minutes.^15^ In this study, we assumed that once water reaches boiling temperature (calculated as described above), it is boiled at 100°C for 1 minute, based on CDC and WHO recommendations, to reach the assumed LRV. The influence of elevation on boiling temperature was not considered in this analysis, but this would impact the temperature and thus the length of time needed for boiling for a given LRV.

Once the energy demand was estimated, an air pollution box model^61^ was used to calculate the kitchen PM_2.5_ concentration (µg m^-3^) over 24 hours. The time for cooking and water heating is calculated by the energy demand divided by the assumed stove power (Watts) and thermal efficiency (See Supplemental Material, Emission Factors (EF), Stove Power (SP), and Thermal Efficiency (TE) of Stoves). We assumed that cooking and water heating each occurred once per day to produce all daily energy for cooking and water heating. The emissions were calculated using the relevant emission factor (grams PM_2.5_ per kilogram fuel) and heating value of fuel (See Supplemental Material, Heating Value of Fuels). We also assumed an ambient concentration^67^ of 12 µg m^-3^ to which the indoor concentration decays after the cooking event, and a second event (water heating) increased it again. If a single stove did not provide sufficient power to heat the specified daily water and food allowing time for household concentrations to return to ambient levels between stove uses, a second stove is used in the model simultaneously, and in this scenario, we doubled emission rates. Although emission factors vary between stove operation stages (e.g., startup vs steady operation),^68–71^ we assumed a constant average emission factor. This approach may underestimate 24-hour emissions, since it doesn’t reflect multiple starting events which can produce high emissions.^72^ For an example of the 24-hour PM_2.5_ kitchen concentration for cooking and water heating, see Supplemental Material, Example Air Pollution Box Model Run of 24 hour PM_2.5_ Concentration.

To calculate the health burden from IAP, we adapted the approach of the Household Air Pollution Intervention Tool (HAPIT).^41,67^ Personal exposure is estimated by multiplying the modeled 24-hour PM_2.5_ kitchen concentration by an estimated ratio of personal exposures to kitchen concentrations^42,73,74^ with separate ratios applied for the cook, non-cook, and children under 5 of a household (See Supplemental Material, Personal Exposure to Concentration Ratios).

The burden of disease attributable to household PM_2.5_ pollution is calculated for lung cancer (LC), ischemic heart disease (IHD), stroke, acute lower respiratory infection (ALRI), and chronic obstructive pulmonary disease (COPD) using 2019 background disease data, deaths & DALYS ^75,76^ for Uganda and Vietnam, respectively, and disease-specific integrated exposure response functions.^77^ While there is no clear safe level of PM_2.5_ exposure,^77^ we use the suggested distribution of 5.8-8.8 μg/m^3^ as a counterfactual ‘no effect’ level.^74^

The relative risk and the existing fraction of each country’s population exposed to solid fuels (i.e., fraction exposed equals the percent solid fuel users for each country) is used to calculate the attributable fraction (AF) (Equation 3).^67^ In calculating the attributable fraction in the model, the fraction exposed is country-specific and fixed, but the relative risk varies with air pollution level.

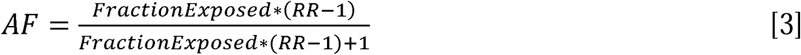

The attributable fraction is multiplied by the DALYs or deaths from a given disease in the country or region of that specific population (given location and age group) to calculate the attributable burden associated with IAP.^41^ To calculate the fraction of DALYs from children under 5, we used the fractions of children in that population and relevant under-5 DALYs for each disease.

### Simulations, Analysis, Statistical Tests, & Sensitivity

When comparing IAP and drinking water DALYs, we define ‘net DALYs’ as the increase in IAP DALYs minus the decrease in drinking water DALYs resulting from boiling drinking water. Positive net DALYs means that the IAP DALYs increase is greater than the water DALY decrease, indicating a net increase in disease burden. Negative net DALYs means the water DALYs decrease is greater than the IAP increase, indicating a net health benefit.

We conducted Monte Carlo simulations to capture the influence of variability and uncertainty in the inputs. Each simulation draws from distributions for parameters of a stove and fuel type, and water boiling effectiveness and is run 1000 times in R.^78^ The mean, standard deviation, and 95% confidence intervals of model output are reported. Additionally, to understand the variation in the outputs, the coefficient of variation (COV) (standard deviation over mean) was calculated. Environmental parameters are often log-normally distributed,^79^ so we used the Shapiro-Wilk test to test the normality of our resulting distributions of drinking water and IAP DALYs before making statistical comparisons between different scenarios. If an output distribution was log-normal, it was log-transformed for hypothesis testing. We used the t-test to compare the risks across the different scenarios (i.e., different stove types or water boiling scenarios). P-values less than 0.05 were considered statistically significant.

A sensitivity analysis was conducted for the IAP and drinking water QMRA models to identify the specific impact of individual input variables on model output. Each input parameter was individually evaluated by varying the assigned value between a minimum and maximum determined based on an assessment of the variability or uncertainty of the parameter from our literature review (See Supplemental Material, Parameters Varied in the Sensitivity Analysis for Indoor Air Pollution and Parameters Used in the Sensitivity Analysis of the Water Risk Module). Input parameters were then ranked in order of their influence on output values by taking the ratio of the output values for input at its minimum and maximum values.^80^

## Results

### DALYs from Indoor Air Pollution

The simulations with the lowest to highest 24 h average PM_2.5_ kitchen concentrations (for cooking and water heating scenarios) were electric, LPG, gasifier, charcoal, improved wood, and traditional wood, with values ranging from 12 µg/m^3^ (for the electric stove) to 5587 µg/m^3^. The average 24-hour PM_2.5_ concentration for the ‘worst-case’, traditional wood stove scenario was lower for water heating alone (2109 µg/m^3^) than for cooking alone (3490 µg/m^3^), while the concentration associated with both activities together was essentially the same as the sum of the two activities considered separately.

Figure 2 shows the DALYs calculated from these PM_2.5_ values for the different stove and use scenarios for each country. Though the total DALYs per 10,000 people were similar between the two countries, the number of IAP DALYs associated with under-5s were higher in Uganda compared to Vietnam, with the under-5s DALYs in Uganda making up 48% to 64% (depending on scenario) of total, versus under-5s DALYs in Vietnam making up 5% to 8% of total DALYs. DALYs significantly decrease as stove type shifts from traditional wood to LPG stoves, with 95.3%, 96.9%, 97.1%, and 99.9% decrease in DALYs for Uganda over-5’s, Uganda under-5s, Vietnam over-5’s, and Vietnam under-5s respectively, eliminating almost all the DALYs associated with IAP. For the electric stove, since it is assumed that the electric stove contributes no additional PM_2.5_, there was a 100% decrease. There are only small changes in DALYs when a traditional wood stove is replaced with an improved wood stove. For example, DALYs decrease by 1.0% (for Uganda children) to 11.2% (Vietnam over-5’s) for water heating and cooking when switching from traditional wood stove to improved stove, indicating the limited potential for health impacts from replacing traditional stoves with another basic wood stove.

**Figure 2.**
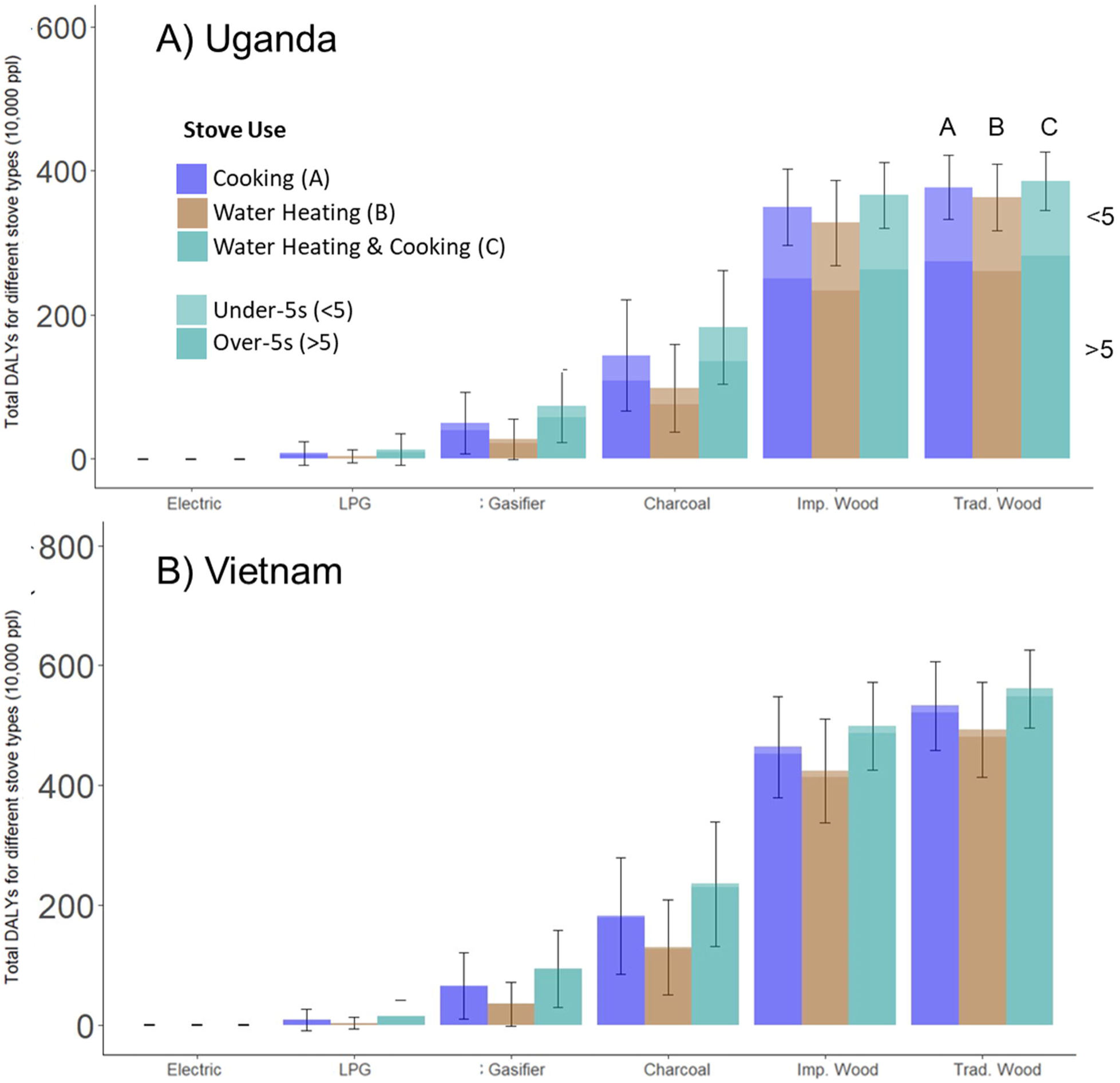
Mean DALYs from indoor air pollution for Different Stove and Use Scenarios for Uganda (A) and Vietnam (B). Error bars show+/- one standard deviation. Baseline is no stove use. DALYs calculated for scenarios of cooking only, water boiling, and water boiling plus cooking. Under-5s refers to children less than 5 years of age and over-5s as all other age groups combined.

In contrast to the additive nature of the PM_2.5_ concentration when aggregating water heating and cooking, DALYs are not additive due to the log-linear nature of the dose response curve.^81,82^ As a result, the DALYs associated with ‘only cooking’ and ‘water heating and cooking’ are similar (e.g., mean of 377 and 386 per 10,000 people, respectively, for traditional wood stove in Uganda for all ages), suggesting that the additional exposure from boiling adds a minimal increment to the IAP impact. However, results in Figure 2 show that the relative increment (fractional increase in DALYs from adding boiling) is slightly larger for cleaner cooking options.

### DALYs from Water Contamination

The total DALYs from drinking water for Uganda and Vietnam were similar, but a larger share of the DALYs came from children under 5 in Uganda (21-27% compared to 12-14% in Vietnam) because of the higher number of children per household in Uganda. DALYs from drinking water were a strong function of untreated water quality and boiling effectiveness. For example, for over-5s in Uganda, and low-risk water, health impacts ranged from 51 DALYs per 10,000 people per year for lab-level boiling to 210 DALYs per 10,000 people per year from ineffective boiling, compared to 220 DALYs without boiling. For high-risk water, DALYs ranged from 59 per 10,000 people per year for lab-level boiling to 1137 per 10,000 people per year for ineffective boiling, compared with 1168 per 10,000 people per year without boiling. Using lab-level boiling compared to untreated water greatly reduced adult DALYs in both Uganda and Vietnam. For Ugandan children under 5, lab-level boiling decreased DALYs by 94%, 88% and 72% for high-, medium-, and low-risk water, respectively, compared to ineffective boiling. Relative reductions in DALYs for children under 5 in Vietnam were similar.

### Comparison of water and air pollution DALYs

In most of our scenarios, the benefits of clean water outweighed the impacts of IAP from boiling that water. Figure 3 reports the changes in DALYs for water and IAP exposures under various boiling and stove scenarios. The higher fraction of DALYs for children under 5 years in Uganda for both water and air reflect their higher proportion in the population. Additionally, the relative contribution of the under-5s category was much greater for the IAP risk (driven by ALRI) than for water.

**Figure 3.**
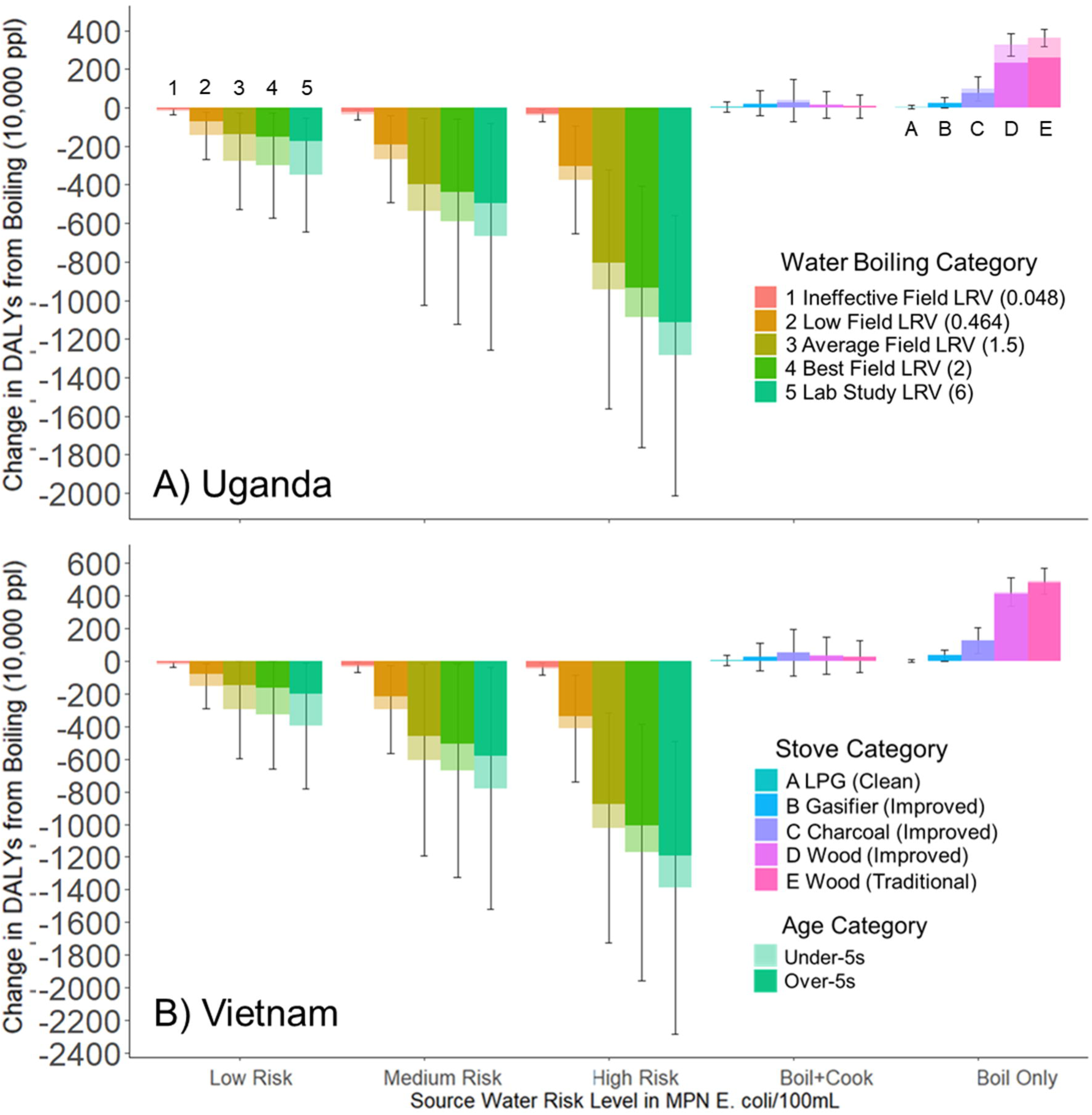
Mean change in DALYs from boiling drinking water with different stove types for under-5s and over-5s in Uganda (A) and Vietnam (BJ for different stove types and use (assuming households either previously cooked and did not cook), and for different water source qualities and boiling effectiveness. The error bars show +/- one standard deviation from Monte Carlo simulations.

All source water categories boiled with lab-level effectiveness (LRV=6) had reductions in waterborne DALYs greater than increases in IAP DALYs for all stove types and stove use scenarios (water heating without cooking, and water heating with cooking). For the scenario of a household already cooking, and beginning to also boil their water, we found that if the source water is boiled with at least the “low field” effectiveness (LRV of 0.5 or greater), the decrease in drinking water DALYs from boiling was greater than the increase in IAP DALYs for all scenarios of water quality, water boiling, and stove types. We observe a net reduction if a household is already cooking and the water is boiled with at least low field effectiveness for all water risk categories and all stove types. If a household is not already cooking, there is a net benefit if medium- or high-risk water is boiled with at least average field effectiveness for all stove types.

### Log-removal rates needed for health benefits from water boiling

As another way to examine net benefits of boiling, the model was also used to determine ‘break-even’ points where increases in IAP DALYs are equal to the associated decrease in drinking water DALYs. Figure 4 shows the reduction in drinking water DALYs for over-5s in Uganda plotted against LRVs for low-risk, medium-risk, and high-risk water qualities. The increase in IAP DALYs is also shown for two stoves, traditional and LPG, and for two different use scenarios, only water heating and water heating plus cooking. Where these two sets of curves cross can be considered the ‘break-even’ point, or the minimum LRV required to provide a net reduction in DALYs for a given stove scenario. High-risk water has a significant decrease in DALYs even with low LRVs. The large decrease in pathogen exposures even with low LRVs results in large health benefits. For the case of over-5s in Uganda, the intersection of the curves and IAP lines show that when cooking is already taking place in a home (cooking compared to cooking plus water heating), boiling drinking water with an LRV of 0.18 or greater results in a net reduction in DALYs for all stove types and source water types, including the dirtiest stove (traditional wood stove) and low-risk water. In the case that a household is not cooking, but starts boiling water, LRVs of 0.2, 0.3, and 1 are needed for high-, medium-, and low-risk water, respectively, to offset DALYs from IAP. The LRV cutoff needed to achieve net health benefits increases as water quality improves (for example, an LRV of 0.2 is needed for high-risk water compared to an LRV of 1 for low-risk water).

**Figure 4.**
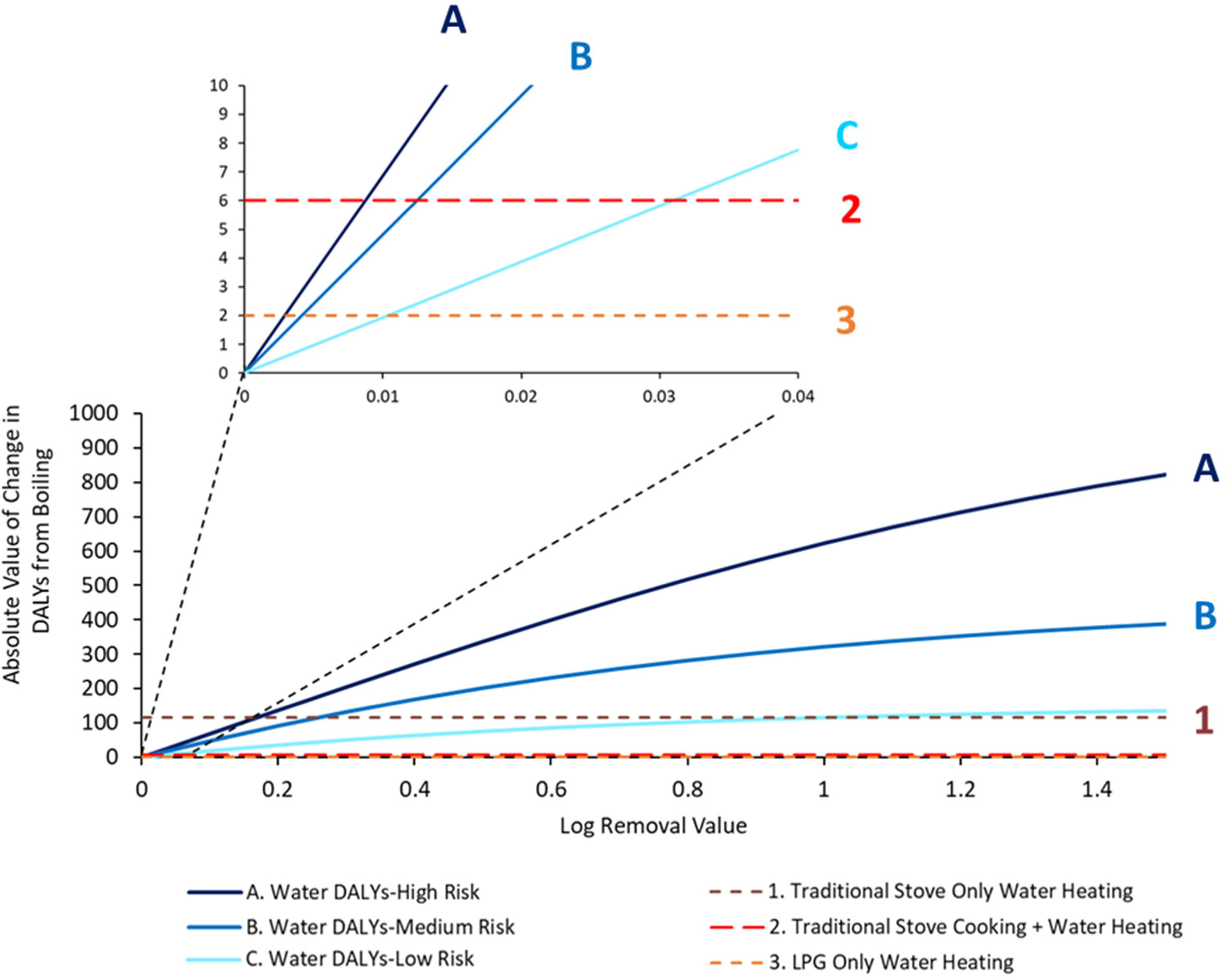
Absolute value of change in DALYs from boiling in water (reduction in Drinking Water DALYs and increase in /AP DALYs) versus boiling log removal value (LRV) for homes in Uganda, consideringover-5s only. The increase in DALYs, compared to baselines of both cooking and not cooking, are shown for LPG and traditional stoves. Water risk levels of low, medium, and high E. coli levels are shown. Inset shows a zoomed in version of the large panel with same axes to show tradeojfs at low LRVs.

### Sensitivity Analysis

Estimated reductions in DALYs from water boiling were most sensitive to following parameters (listed from greatest to lowest sensitivity): source water *E. coli* levels, the assumed ratios of *Cryptosporidium*, rotavirus, and *Campylobacter* to *E. coli*, age at death, dose-response parameters used in the QMRA, the LRVs, and the water volume ingested. For IAP DALYs, the input parameters with greatest influence (ranked from greatest to least) were: stove emission factor, household air exchange rate, fuel heating value, and room volume. For the sensitivity of the QMRA dose-response parameters, we varied the parameters for each of the three pathogens, and the level of risk per ingested dose. Of the three pathogens considered in the QMRA, rotavirus has the highest risk per dose of all the pathogens, and as a result, rotavirus exposure had the largest influence on QMRA DALYs (See Supplemental Material, Sensitivity Analysis Showing Net Difference in Total DALYs, Sensitivity Analysis for Water DALYs, and Sensitivity Analysis for Water DALYs: Pathogen Specific Parameters).

## Discussion

In this study, we find that in most scenarios (even at low LRVs and with high-emitting cookstoves), boiling medium- and high-risk drinking water results in a net decrease in total DALYs. It is estimated that 1.1 billion people drink water that is of at least moderate risk,^83^ so boiling would likely benefit these 1.1 billion people. Additionally, though 89% of the world’s population uses an improved drinking source, such as household connections, public standpipes, boreholes, and protected wells, springs, and rainwater collection,^84^ and the odds of contamination are lower for so-called improved sources,^85^ 10% of improved sources are still considered high risk.^83^ Thus, boiling could benefit even those using improved sources. However, in situations in which a high emitting stove is used to boil water, and cooking is not already taking place in a home or cooking is done on a clean stove, boiling could have a net negative impact, showing the importance of considering risks from both water and air jointly.

Adoption of a new technology or practice is a major challenge for both water and stove interventions and will impact the generalizability of our model results. For example, in an intervention study in Rwanda, use of water filters and improved stoves was measured by self-report and spot-check observations, and though most household used the water filter, the majority continued to use traditional stoves.^86^ In our model, switching from traditional wood and improved stoves to LPG or electric stoves resulted in 99% reductions of PM_2.5_ and 96% reductions in the IAP DALYs, showing the huge potential for benefits from clean stoves, including for boiling. However, there are still many limitations to adopting LPG, including cost^87^ and social and cultural perceptions.^88,89^ Additionally, households often combine different stove and fuel types, known as “stove stacking”,^89–91^ limiting the health benefits of a stove intervention. Traditional stoves continue to be used due to their additional benefits, including heating the living space, lighting the home, heating water for bathing and washing, drying, smoking food, getting rid of waste, keeping insects and animals away, and social gatherings.^92^

This can be especially relevant for water boiling, as previous studies have observed that households continued use of fuelwood to heat water after LPG and electricity are available,^92,93^ possibly due to the cost^94^ of such an energy intensive task. Despite these challenges, the use of polluting stoves such as biomass has continued to decrease, from 53% using polluting stoves in 1990 to 36% in 2020.^5^ However, there have been increases in charcoal use in many areas,^5^ which our model suggests only moderately reduces PM_2.5_ relative to traditional wood use.

Boiling effectiveness, which varies widely, is key in determining whether boiling has a net health benefit. None of the field studies of boiling used for this study has observed ‘lab-quality’ LRVs, and many field studies reported very low, even negative LRVs for boiling.^16^ Measured LRVs comparing post treatment to untreated water (from the source) are influenced by source water quality before it is boiled, post-boiling storage, dipping, and other sources of recontamination.^15–17^ Thus various factors can affect the end water quality despite treatment, limiting the potential benefits of boiling and resulting in a lower effective LRV. Our results show that boiling is most beneficial in terms of a net reduction in scenarios with high risk water; however, homes with high risk drinking water may have the highest risk of recontamination due to poor sanitation practices.^95^ Therefore, it is important to maintain improvements in water quality post boiling if water is not immediately consumed.^15,16,96^

Benefits from increasing LRVs from different water treatment methods strongly depended on compliance of use^97^ and full adoption of the intervention. It has been found that a few days of untreated water consumption after drinking treated water can completely negate the annual health benefits of drinking treated water.^98^ From the literature review conducted for this study on boiling LRVs, societies and cultures with higher prevalence of boiling (e.g., Vietnam) have the highest LRVs.^15^ In countries like Zambia where boiling is not prevalent,^13^ and boiling is promoted as an intervention, the LRVs tend to be much lower.^16^ This suggests that introducing boiling as a water treatment method in a community that does not have a history of boiling presents additional challenges, and likely reduces the chances of effective boiling and a net decrease in DALYs.^16^

One notable alternative to boiling is chlorination, which is cheap, effective, provides residual protection,^99,100^ and does not impact air quality. Chlorination has been shown to reduce the risk of child diarrhea and reduce risk of stored water contamination by *E. coli.*^101^ Residual protection could result in higher real-world LRVs. Unlike boiling, chlorination can be applied in several different ways, such as at the source and before drinking.^102^ The use of chlorination at the water source (such as chlorinating a pump or water source) also eliminates the need for behavior change practices in the home. Chlorination has been observed to be more effective than boiling for reducing *E. coli* concentrations in field settings, possibly due to insufficient heating during boiling and recontamination after treatment.^99^ The residual disinfection in household storage provided by chlorination provides a notable advantage over boiling. However, there are several limitations related to chlorination, including challenges with the supply chain^103^ and managing its use (including frequency and amount of added chlorine).^104^ There are also limitations in the effectiveness of chlorination. One study found that combining chlorination with boiling did not lower *E. coli* contamination in stored water or lower risk of diarrhea compared with boiling alone.^99^ There are conflicting data on the effectiveness of boiling versus chlorination. In one laboratory-based study, boiling was more effective in inactivating *E. coli* and environmental bacteria compared to chlorination and pasteurization.^105^ Additionally, chlorine has limited effectiveness in inactivating viruses and protozoa.^106^ However, in a study of a commercial chlorination product in drinking water, chlorination was found to be associated with lower *E. coli* contamination and diarrhea rates in children under 5 years than boiling.^99^ Other downsides of chlorination include production of byproducts, taste/odor concerns which can lead to low adoption, and cost, which is low, but remains a consideration.^106^ Both boiling and chlorination have tradeoffs; however, our study suggests that the minimal IAP impacts associated with boiling should likely not the central factor in deciding whether boiling or chlorination is the best option for a household.

The cost of stoves and fuels were not considered in this study. However, cost is a major factor in whether a household adopts drinking water treatment^107^ or improved stoves.^108,109^ Cost-benefit analyses have been conducted for cookstoves. For example, one study compared capital, operation and maintenance, and environmental costs to benefits such as health, supply, ease of operation, and safety and found the lowest cost-benefit ratio for traditional biomass cookstoves (lower than improved biomass), second lowest for dung, and the highest for biogas, charcoal, and LPG.^110^ Another study including only improved stoves (including LPG and improved cookstoves), had the lowest cost to benefit ratio for LPG and highest for improved biomass stoves.^111^ While study assumptions and approaches vary, these findings highlight the potential challenges facing users in switching to improved and cleaner stoves. A cost benefit analysis on water treatment showed the benefits of the filters outweighed the costs only when including aesthetic benefits, not health benefits alone.^112^ A cost-benefit analysis for a combined cookstove and water treatment intervention (an improved stove and water filter) in Rwanda showed that benefits from fuelwood savings, time savings, and environmental and health improvements outweighed the cost.^33^ However, fuel savings among new water filter users were found only for households who previously boiled their water, not for households which had not treated their water previously.^33^ A cost benefit analysis comparing boiling with various stove/fuel options could expand on our findings and add another dimension to the tradeoffs we explore. Many clean stoves (e.g. electric, LPG) may be currently cost-prohibitive in low-income settings.^113^ Additionally, countries with high adoption of LPG due to subsidies (e.g. Ecuador) retain substantial solid fuel use.^114^ In order to implement effective IAP and WASH interventions, cost effective solutions for both should be prioritized.

### Strengths and Limitations

Our model is designed to give insights into possible tradeoffs when addressing multiple environmental health risks. Because the focus of this study was to develop a model framework applicable to many countries and scenarios with a focus on exploring health tradeoffs rather than detailed not contextual differences, this study used many of the same values (e.g., emission factors, fuel efficiency, water *E. coli* levels) for Uganda and Vietnam. Some inputs were country specific, such as demographic parameters. Use of context-specific data on parameters such as stove type or source water quality would enable a more accurate and detailed analysis of options. Some such data are available for our case study countries, such as stove emission factors^115,116^ and source drinking water quality^117,118^ specific to Vietnam and Uganda, water boiling effectiveness in Vietnam,^15^ and studies of water boiling habits in Uganda.^119^ Additionally, some related studies have been done which could be points of comparison, including indoor air pollution levels in Vietnam^120^ and Uganda.^121^ Our study could be refined by using available data specific to Uganda and Vietnam. However, data for the same settings that covers the multiple dimensions in our analysis (e.g. boiling and air quality data for individual, or nationally representative, settings in a given country) is not available. Therefore, targeted field study of water boiling, pathogen levels in pre- and post-treated water, and indoor air pollution concentrations could be beneficial to evaluate boiling in a real-world setting.

Various assumptions used in our IAP modeling could be addressed with more specific input data or targeted measurements. For example, some of the stoves used in our model had poor efficiency, low stove power and low fuel heating values, and thus they are challenged to meet a households water boiling needs. For these stoves, it is assumed that the stove is used up to 24 hours for the household’s cooking needs (as opposed to using multiple stoves for a shorter period). This simplifies the model and avoids assumptions about stove use timing, but is not realistic. The emission factors used for the stoves are averages even though emission factors change throughout the combustion process.^68,69,71^ However, limited data are available on phase-specific emission factors, which necessitated the use of averages. Emission rates are assumed to be the same for cooking and water heating for this study, but are likely different, since it has been shown that varying cooking styles (e.g., frying vs boiling) are associated with different emission rates.^122^ However, lacking cooking- and boiling-specific emissions for Uganda and Vietnam, averages values were used as a reasonable assumption. Another important simplification is our assumption that a single stove was used for all household uses, ignoring stove stacking.^89,123^

The box model used to calculate 24-hour PM_2.5_ has been found to overestimate concentrations,^61,124,125^ and so modeled exposures are likely biased high. Depending on where the actual PM_2.5_ exposure falls on the dose response curve, this means we could be over- or under-estimating the net change in DALYs due to the added activity for boiling. For example, if the PM_2.5_ with and without boiling falls above where the curves ‘plateau’, the difference in DALYs we estimated could be smaller than in reality.

Our modeled values can be compared with past published field measurements. We estimated mean 24 h PM_2.5_ concentrations ranging from 1797 µg/m^3^ to 5212 µg/m^3^, 721 µg/m^3^ to 2068 µg/m^3^, and 14 µg/m^3^ to 16 µg/m^3^e for traditional wood, improved wood and LPG stove use cases, respectively. In a cross-sectional study in India in four states, the measured mean 24-hour PM_2.5_ concentrations were 590 µg/m^3^ in the kitchen for a traditional wood stove, and 179 µg/m^3^ in the kitchen for LPG.^126^ In another study in India of an improved stoves intervention, 24 h PM_2.5_ for LPG stoves ranged from 70 to 103 µg/m^3^, and was around 500 µg/m^3^ for households using a traditional three stone fire.^127^ A study in Nepal found 24 h PM_2.5_ of 80 µg/m^3^ for electric stoves and 630 µg/m^3^ for traditional mud wood stoves.^128^ These findings suggest that we both overestimate the impacts of traditional cooking and the benefits of ‘realistic’ clean stove adoption in many settings. For example, our model calculated substantially lower values for LPG-using households compared to the field studies, suggesting we may overestimate the benefits of LPG use. Our modeled values for traditional wood stoves were higher than the field studies. Our estimated 24 h PM_2.5_ concentrations for improved biomass stoves are similar to those observed for traditional wood stoves in several field studies, suggesting a bias in model parameters used as defaults. The is potentially at least partly due to stove stacking in the field studies, since many study households used multiple stoves.^128^

That modeled PM_2.5_ values for traditional stoves are much higher than typically observed in field studies suggests that assumptions in IAP model (which is used in WHO assessments) lead to estimates substantially higher than reality. However in our application, such a high bias in model results is likely conservative (overestimates IAP impacts while boiling with traditional biomass), and further emphasizes our finding that water boiling typically has a net benefit, even if using a very high-emitting stove. The relative differences we found between exposures associated with different stoves is fairly representative of real-world observations. For example, a 48% reduction in cooking area PM_2.5_ was measured during an improved wood stove intervention study in Rwanda,^86^ relative to our average modeled reductions of 62% for PM_2.5_ concentrations for a similar scenario (3490 µg/m^3^ and 1334 µg/m^3^ for traditional and improved stoves, respectively). However, as acknowledged above, the log-linearity of the PM_2.5_ dose-response curves means that depending on the pre-/post-boiling combination, the modeling could over- or underestimate the DALYs associated with the addition of boiling. Future work can explore this using a re-calibrated model, or representative field observations.

Our results can also be compared with previous indoor air quality modeling studies. The study presenting the indoor air pollution box model used here estimated kitchen concentrations during cooking, with 24-hr averages ranging from 15 ug/m^3^ for LPG to 1975 ug/m^3^ for traditional stoves.^61^ The estimated PM_2.5_ concentrations for wood stoves from our study using the model are higher, due to different emission factor, stove power, and thermal efficiency inputs for the traditional stoves. A study in China used this air pollution box model in a more sophisticated way with substantially more household data to refine assumptions. This study used inputs from past field studies in China, assigned a ventilation index based on observations of cooking area ventilation (presumably via a chimney or other active ventilation), and information on cooking and living area location to scale the model air exchange rate. This study reported average modeled kitchen PM_2.5_ concentrations of 79 µg/m^3^ for households using biomass to boil water, with the three homes with poor ventilation having an average of 148.3 µg/m^3^.^19^ Our estimated PM_2.5_ from water heating was significantly higher, likely due to the inclusion of a ventilation parameter and stove emission factors specific to China,^19,129^ so a direct comparison is likely not warranted. However, this study provides an example of how IAP modeling can be refined for future studies of specific locations.

Environmental factors outside the home may also have an important influence on IAP not addressed in our model. For example, although ambient air pollution levels in LMICs vary widely across countries and between urban and rural areas,^81,130^ we assumed a fixed background ambient PM_2.5_ concentration of 12 µg/m^3^. If actual ambient air pollution levels are higher, PM_2.5_ exposure levels are shifted higher on the dose response curves, resulting in a smaller additional risk from the increment of PM_2.5_ from boiling water. We also considered only cooking and water heating, and did not consider space heating, even though many countries in the world where boiling is common are located in cold regions.^11,13^ Additionally, it is assumed that boiling is only used for drinking water, not for making tea or bathing. However, if a household is burning fuel for space heating or other reasons, and boiling can be done simultaneously, no additional IAP DALYs could be incurred to boil water.

Several simplifying assumptions made in the water module also influence our results. For example, though we separated by population over and under 5, we assumed a constant probability of death for all ages. However, this likely leads to an underestimation of the years of life lost due to death from diarrhea since children have the greatest risk of death and disease from diarrheal diseases.^131^ Children do not have a fully developed immunity which makes them more susceptible to diarrheal illnesses, which is not accounted for in the dose-response curves used in the modeling. Therefore, if child-specific dose-response curves were available and used, the benefits of boiling would likely be greater.

We estimated pathogen levels based on fecal indicator bacteria (i.e., *E. coli)* levels, which are imperfectly correlated with the selected pathogens.^132^ Further, pathogens present in drinking water vary widely by country and across seasons.^133^ We expect that the results would vary widely based on the pathogens selected, as has been found in previous studies.^134^ Our results were highly sensitive to parameters related to pathogen risk. Thus, pathogen-specific water quality data would be helpful to characterize risks associated with pre- and post-boiling water. Limited data exist, but new methods (with their own associated limitations) are expanding the potential for pathogen specific data, such as the use of TaqMan array cards for environmental samples.^135^ Additionally, water quality in LMICS can be highly temporally variable, with spikes in contamination, and intermittent exposure to contaminated drinking water can negate the benefits of improved water access.

DALYs are used for comparative risk assessment, including environmental risks such as IAP and water contamination. DALYs allow comparison across risk categories, and are the common metric used in the Global Burden of Disease estimates.^136^ Estimates for DALYs from air pollution and water are calculated using very different approaches, which we adopted for this study. Additionally, though DALYs account for the years of life lost and years lived with disability, they don’t perfectly capture the acute versus chronic nature of disease, which is an important difference between IAP and water risks. Using these different methods to compare different risks and outcomes is an inherent limitation in the comparative risk assessment approach. However, we took this approach as it is the most widely used approach to compare diverse risk factors. Future work can examine other methods to calculate and compare the risks from drinking water and IAP, including field studies to provide context-specific findings to compare against the broad insights from this scoping analysis.

## Conclusions

To our knowledge, this is the first study to compare water and air pollution DALYs for scenarios of household cooking and water boiling to determine if boiling water has a net benefit regardless of stove and fuel type. Our model identified certain scenarios that, if risks for water and air are considered separately, could suggest DALY reductions but that due to combined effects ultimately results in net positive DALYs. For example, if a household that does not cook their food is encouraged to boil water on a wood stove, they would experience increased air pollution exposure that may outweigh benefits of ineffectual water boiling. However, this scenario is likely an exception, and our modeling suggests that boiling even at low log-removal rates has net health benefits for medium- and high-risk source water, even if using relatively high-emitting stove. In contrast, when treating low-risk source water, boiling effectiveness and cookstove type influence whether net health benefits result from the practice. We recommend further field investigations to jointly assess health effects associated with contaminated drinking water and indoor air pollution and the development and evaluation of interventions to mitigate both exposures. Due to uncertainties and assumptions in the model, we recommend that country or context-specific inputs, and ideally studies of boiling and indoor air pollution, with empirical measurements of water and air quality, be conducted to improve understanding of the health risks and tradeoffs and evaluate our model. Specifically, more information on household-specific parameters, including pathogen contamination in water, stove types, and cooking and boiling energy requirements, and linking emissions to indoor air pollution and exposure, could improve the model. Future efforts should empirically measure air pollution concentrations (to estimate health impacts) and diarrhea during field studies of different boiling methods and stove types, to both provide better input parameters and improve our ability to model risk tradeoffs.

## Data Availability

https://datadryad.org/stash/share/t6EFh-eWi7ORidNbrcWb1BV9yKQ6PyClW_9XZYDJhGE

## Acknowledgements

Special thanks to Sean Daly for his suggestions and feedback on this project. Thank you to Cheryl Weyant for providing feedback on the writing. We would like to dedicate this paper to the memory of Haley Foard, a North Carolina State University undergraduate researcher who contributed to the initial literature review and research in the initial phase of the project during Summer 2020.

## Appendices

## Appendix 1: Key Assumptions for Indoor Air Pollution DALYs Calculation

- Use of an indoor air pollution box model ^125^
- Kitchen characteristics assumed to be the same in all scenario, with volume of 30 (sd 15) m^3^ and air exchange rate of 25 hr^-1^ (sd 15) ^61^.
- Only considered disease burdens from 5 outcomes: Stroke, ALRI, IHD, Lung Cancer, and COPD.
- Used Country Level Disease Burdens for a single year, 2019.
- Assumed the same exposure to concentration ratio regardless of location.
- Used a single average PM_2.5_ emission factor over the entire burning process.
- Assumed all cooking and all water heating each occurred one time per day.
- Does not consider space heating.
- Assumes ambient air pollution level of 12 µg/m^3.^
- Used integrated exposure response functions to calculate risk for each disease outcome ^77^
- Integrated exposure function assumes 1) exposure to PM_2.5_ from diverse combustion sources is associated with increased mortality; 2) they are a function of PM_2.5_ mass inhaled concentration across all combustion particle sources and composition; 3) there is no interaction between different exposure types^77^
- Used counter-factual exposure value of a distribution of around 7 μg/m^3^ for the IAP exposure response model.^77^

## Appendix 2: Key Assumptions for Drinking Water DALYs Calculation

- Characterized risk using three indicator pathogens.
- Assumed disease burden is based on calculating DALYs from *E. Coli*.
- Assumed constant ratios for converting disease pathogens to *E. Coli*.
- Assume three categories of water risk level.
- Neglected post boiling recontamination.
- Assumed boiling takes place one time per day.
- Assumed all ages are at the same risk from the disease and uses same calculations for all ages.
- Age difference is the only demographic difference.

