## Supplemental Tables and Figures for "Do the Health Benefits of Boiling Drinking Water Outweigh the Negative Impacts of Increased Indoor Air Pollution Exposure?"

Angela Harris:

3213 Fitts-Woolard Hall

915 Partners Way

Raleigh, NC 27607

Andrew Grieshop:

3165 Fitts-Woolard Hall

915 Partners Way

Raleigh, NC 27607

Conflict of Interest: The authors declare they have nothing to disclose.

### Tables

Table S 1 Parameters Needed for Model

| IAP Model | QMRA Model | Demographics |
| --- | --- | --- |
| Drinking water consumed | *E. coli* level in drinking water | Number of people in household |
| Fuel emission factor | Dose response parameters of pathogens | Number of cooks, noncooks, and children per household |
| Fuel Heating value | Burden of disease for each pathogen | Fraction of population which is under 5 |
| Stove power | probability of illness | Life Expectancy men |
| Thermal Efficiency | percent susceptible | Life Expectancy women |
|  | boiling effectiveness | Percent solid fuel users |

Table S 2 Household Demographics for Vietnam and Uganda

|  | Vietnam | Uganda | Source |
| --- | --- | --- | --- |
| Average Household Size | 5 | 5 | (United Nations, 2017) |
| Life Expectancy Women | 79.2 | 69.2 | (Institute for Health Metrics and Evaluation, 2021) |
| Life Expectancy Men | 70 | 62.3 | (Institute for Health Metrics and Evaluation, 2021) |
| Adults Per Household | 4.6 | 4 | (Pillarisetti et al., 2015) |
| Children Under 5 Per household | 0.4 | 1 | (Pillarisetti et al., 2015) |
| Total Country Population | 96362928 | 41117856 | (Institute for Health Metrics and Evaluation, 2021) |
| Cooks per household | 1 | 1 | (Pillarisetti et al., 2015) |
| Non Cooks over 5 per household | 3.6 | 3 | (Pillarisetti et al., 2015) |
| Non Cooks under 5 per household | 0.4 | 1 | (Pillarisetti et al., 2015) |
| Percent Solid Fuel Use | 49% | 95% | (Pillarisetti et al., 2015) |

Table S 3 Ratio of E. coli to Campylobacter, Cryptosporidium, and Rotavirus

| *E. coli*: pathogen | Ratio*^a^* | Water Source Used in Study | Ratio used in Calculation | Citation |
| --- | --- | --- | --- | --- |
| E. coli: Campylobacter | 1:0.42 | Filtered surface water (pond) | 1:0.66 | (Kundu et al., 2018; Machdar et al., 2013) |
| E. coli: Campylobacter | 1:0.9 | Ozonated Water | 1:0.66 | (Kundu et al., 2018; Machdar et al., 2013) |
| E. coli: Campylobacter | 1: 8.89x10^-3^ mean, st. dev 1.33 | Sewage |  | (Bivins et al., 2017) |
| E. coli: Rotavirus | 1:10^-5^-10^-6^ | Waste Water | 1:5*10^-6^ | (Machdar et al., 2013) |
| E. coli: protozoa (Used as estimate for Cryptosporidium) | 1:(10^-5^-10^-7^) | Waste water | 1:10^-6^ | (Kundu et al., 2018; Machdar et al., 2013) |

*^a^* Pathogens Distribution is uniform if only mean is provided, and lognormal if mean and standard deviation are provided. References: Machdar et al., 2013; Murphy et al., 2016; Uprety et al., 2020.. For our model, if multiple ratios available, the average value was used (1:0.66 for Campylobacter, 1:5*10^-6^ for rotavirus, and 1:10^-6^ for protozoa)

Table S 4 Log Reduction Values of E. coli, TTC, and FC

| Country | E. coli*^a^* | TTC | FC | Citation | Value used in presented results |
| --- | --- | --- | --- | --- | --- |
| Laboratory (1 minute, 75^o^C) | 6.00 |  |  | (Sanciolo et al., 2015) | Yes |
| Tanzania 1 |  | 2.50 |  | (Mohamed et al., 2016) | No |
| Tanzania 2 |  | 2.19 |  | (Mohamed et al., 2016) | No |
| India |  |  | 2.02 | (Clasen & Rosa, 2017) | No |
| Cambodia surface water | 2.00 |  |  | (Sobsey & Brown, 2012) | Yes |
| Vietnam <2 hr |  | 1.95 |  | (Clasen et al., 2008) | No |
| Cambodia rain | 1.90 |  |  | (Sobsey & Brown, 2012) | No |
| Cambodia uncovered | 1.90 |  |  | (Sobsey & Brown, 2012) | No |
| Cambodia pour | 1.90 |  |  | (Sobsey & Brown, 2012) | No |
| Vietnam flask |  | 1.89 |  | (Clasen et al., 2008) | No |
| Indonesia |  | 1.88 |  | (Sodha et al., 2011) | No |
| Vietnam 2-4hr |  | 1.81 |  | (Clasen et al., 2008) | No |
| Cambodia Covered | 1.80 |  |  | (Sobsey & Brown, 2012) | No |
| Cambodia Dipping | 1.80 |  |  | (Sobsey & Brown, 2012) | Yes |
| Tanzania 3 |  | 1.50 |  | (Mohamed et al., 2016) | No |
| Cambodia Well | 1.50 |  |  | (Sobsey & Brown, 2012) | Yes |
| Vietnam 4-8 hr |  | 1.45 |  | (Clasen et al., 2008) | No |
| Vietnam 8-16 hr |  | 1.29 |  | (Clasen et al., 2008) | No |
| Vietnam Jug |  | 1.27 |  | (Clasen et al., 2008) | No |
| Vietnam Alum. vessel |  | 1.24 |  | (Clasen et al., 2008) | No |
| Vietnam 16-24 hr |  | 1.10 |  | (Clasen et al., 2008) | No |
| Guatemala |  | 0.88 |  | (Rosa et al., 2010) | No |
| Vietnam >24 hr |  | 0.83 |  | (Clasen et al., 2008) | No |
| China Electric kettle |  | 0.56 |  | (Cohen & Colford, 2017) | No |
| Peru Urban Visit 2 |  | 0.44 |  | (Rosa et al., 2014) | Yes |
| China Pot |  | 0.37 |  | (Cohen & Colford, 2017) | No |
| Peru Urban 3 |  | 0.36 |  | (Rosa et al., 2014) | No |
| Peru Urban 1 |  | 0.14 |  | (Rosa et al., 2014) | No |
| Peru Rural 2 |  | 0.09 |  | (Rosa et al., 2014) | No |
| Peru Rural 3 |  | 0.07 |  | (Rosa et al., 2014) | No |
| Peru Rural 1 |  | 0.06 |  | (Rosa et al., 2014) | Yes |
| Zambia same day |  | -0.17 |  | (Psutka et al., 2011) | No |
| Zambia |  | -0.26 |  | (Psutka et al., 2011) | Yes |

*^a^* All values were converted to E. coli for comparison in the model. The righthand column shows the selected values presented in the results. Table continues over 3 pages.

Table S 5 Effect of Boiling on E. coli, Campylobacter, Cryptosporidium, and Rotavirus

| Organisms | Log Reduction value*^a^* | Temperature | Contact Time (seconds) | Sources |
| --- | --- | --- | --- | --- |
| E. coli | 6 | 75^o^C | 60 | (Sanciolo et al., 2015) |
| Campylobacter | 5 | 63^o^C | 300 | (Sanciolo et al., 2015) |
| Cryptosporidium | 8 | 75^o^C | 60 | (Sanciolo et al., 2015) |
| Rotavirus | 8 | 95^o^C | 60 | (El-Senousy, 2020) |

*^a^* To convert to the pathogen specific LRV, the ratio of the LRV for the given pathogen to *E. coli* was used. These were not distributions but specific parameters.

Table S 6 Dose-Response Parameters for the Selected Pathogens

| Organisms | Dose-Response Parameters | Model | Sources |
| --- | --- | --- | --- |
| Campylobacter | *α*: mean=1.51*10^-1^  sd. Dev: 5.9*10^-2^, N_50_: mean=1.69*10^-3^, sd dev: 2.78*10^-3^, P_Di_=0.3 | β-Poisson | (Bivins et al., 2017; Black et al., 2022; Haas et al., 1999; Machdar et al., 2013; Medema et al., 1996; Westrell, 2004) |
| Rotavirus | *α*: mean=2.48*10^-1^, sd. Dev: 1.46*10^-1^, N_50_: mean =8.16, sd. Dev:6.65, P_Di_=0.51 | β-Poisson | (Bivins et al., 2017; Haas et al., 1999; Medema et al., 1996) |
| Cryptosporidium | r: mean=3.44*10^-1^ sd. Dev = 1.46*10^-1^, P_DI_ = 0.7 | Exponential | (Bivins et al., 2017; Messner et al., 2001; Westrell, 2004) |

Table S 7 Probability of Illness Given Infection & Percent Susceptible

| Pathogen | Probability of Illness Given Infection | Percent Susceptible*^a^* | Citation |
| --- | --- | --- | --- |
| Cryptosporidium | 70% | 100% | (Bivins et al., 2017; Machdar et al., 2013) |
| Campylobacter | 30% | 100% | (Bivins et al., 2017; Machdar et al., 2013) |
| Rotavirus | 50% | 13% | (Bivins et al., 2017; Machdar et al., 2013) |

*^a^* Either based on risk or based on population consuming that water source

Table S 8 Burden of Disease for Each Pathogen

| Pathogen | Outcomes | Severity*^a^* | Duration- Days*^b^* | Duration (Years)-based on lifetime (56 here)*^c^* | DALY per case*^d^* | Likelihood of Outcome*^e^* |
| --- | --- | --- | --- | --- | --- | --- |
| Campylobacter | Gastroenteritis-population | 0.067 | 5.1 | 0.014 | 0.0009 | 94% |
| Campylobacter | Gastroenteritis- when see doctor | 0.39 | 8.4 | 0.023 | 0.009 | 6% |
| Campylobacter | Death from gastroenteritis | 1 |  | Life expectancy from population distribution | - | 0.1% |
| Rotavirus | Mild Diarrhea | 0.1 | 7 | 0.02 | 0.0019 | 86% |
| Rotavirus | Severe Diarrhea | 0.23 | 7 | 0.02 | 0.0044 | 14% |
| Rotavirus | Death from Diarrhea | 1 |  | Life expectancy from population distribution | - | 0.7% |
| Cryptosporidium | Watery diarrhea | 0.067 | 3.4 | 0.009 | 0.0024 | 100% |
| Cryptosporidium | Death | 1 |  | Life expectancy from population distribution | - | 0.4% |

*^a,b,c,d,e^* (Machdar et al., 2013)

Table S 9 Emission Factors (EF), Stove Power (SP) & Thermal Efficiency (TE) of Stoves.

| Stove | Category | EF Mean ± St. Dev. g/kg*^a^* | Stove Power (MJ/hr) | Thermal Efficiency % ± St. Dev. ^5^ | Study |
| --- | --- | --- | --- | --- | --- |
| Traditional 3 stone wood (version 1) | Traditional Wood, Malawi | 7.1 ± 1.3 | 7.05 | 14.8 ± 1.8 | (McCracken & Smith, 1998; Wathore et al., 2017) |
| Jiko | Charcoal | 0.47 | 21.8 | 23.30 ± 0.29 | (Obeng et al., 2017) |
| MiniMoto(PP) | Gasifier | 0.079 | 11.0 | 37.9 | (Champion et al., 2020) |
| Philips (PW) | Gasifier | 0.41 | 16.0 | 36 | (Champion et al., 2020) |
| Chitetezo Mbaula | Improved Wood | 3.6 | 4.02 | 20 | (Jagger et al., 2017) |
| LPG | Clean | 0.053 ± 0.04 | 5.504 | 54.4 ± 1.5 | (Shen et al., 2018) |

*^a^* Distributions are lognormal

Table S 10 Heating Value of Fuels

| Fuel | Heating value (MJ/kg) mean, min, max, ± St. Dev.*^a^* | Distribution | | Country | Study |
| --- | --- | --- | --- | --- | --- |
| Wood | 14.780, 13.500, 15.883 | Triangular | Malawi | | (Brouwer et al., 1996) |
| Charcoal | 35.9 | Point | Zambia | | (Atteridge et al., 2013) |
| Pellet: Peanut Hull | 16.1 ± 0.18 | Normal | Malawi | | (Champion et al., 2020) |
| LPG | 45.780 | Point | NA | | (Panigrahy et al., 2016) |

*^a^* Dry basis given for biomass, lower heating value is used for LPG

Table S11 Personal Exposure to Concentration Ratios

| Person | Women | Young children | Men | Women to Children | Women to Men |
| --- | --- | --- | --- | --- | --- |
| Ratio^8^ | 0.742 | 0.628 | 0.45 | 0.85 | 0.61 |

*^a^* (Pillarisetti et al., 2015)

Table S 12 Parameters Varied in the Sensitivity Analysis for Indoor Air Pollution

| Parameter | Unit | Minimum | Mean | Maximum | Citation |
| --- | --- | --- | --- | --- | --- |
| Air Exchange Rate | Hour^-1 | 1.39 | 10.4 | 24.9 | (McCracken & Smith, 1998; Parajuli et al., 2016; Park & Lee, 2003) |
| Volume | Meter^3 | 3 | 30 | 100 | (Huboyo et al., 2018; Jagger et al., 2017; Johnson et al., 2011) |
| Daily Cooking Energy | MJ | 1.87 | 10.84 | 45.85 | (Johnson et al., 2011) |
| Daily water consumed and boiled | Liters | 0.67 | 2.55 | 6.10 | (Del Razo et al., 2002; Hossain et al., 2013; Kile et al., 2007) |
| Stove Efficiency | Percent | 10 | 14 | 19 | (Obeng et al., 2017; Shen et al., 2018; Wathore et al., 2017) |
| Fuel Emission Factor | g/kg | 3.98 | 7.00 | 11.76 | (Champion et al., 2020; Shen et al., 2018; Weyant et al., 2019) |
| Stove Power | MJ/hour | 0.64 | 4.9 | 34 | (Champion et al., 2020; Obeng et al., 2017; Shen et al., 2018) |
| Fuel Heating Value | MJ/kg | 13.5 | 19.49 | 45.780 | (Brouwer et al., 1996; Champion et al., 2020; Panigrahy et al., 2016) |
| Exposure to concentration ratios (women)- used for all household members in sensitivity analysis | Fraction | 0.33 | 0.35 | 0.74 | (Shupler et al., 2018) |

Table S 13 Parameters Used in the Sensitivity Analysis of the Water Risk Module

| Parameter | Unit | Minimum | Mean | Maximum | Citation |
| --- | --- | --- | --- | --- | --- |
| Age at Death | # | 1 | 66.7 | 90 | (World Health Organization, 2023) |
| *E. coli* level (literature based) | MPN | 9.02*10^-7^ | 1.66*10^-1^ | 116 | (Bivins et al., 2017) |
| Log Reduction | Unitless | -0.208 | 2 | 6 | (Sobsey & Brown, 2012) |
| K value (Cryptosporidium) | Unitless | 7.89*10^-5^ | 2.77*10^-1^ | 8.10 | (Bivins et al., 2017) |
| Ratio *Cryptosporidium* to e. coli | Unitless | 2.28*10^-10^ | 1.126*10^-6^ | 6.28*10^-5^ | (Bivins et al., 2017) |
| Alpha value (*Rotavirus*) | Unitless | 0.0382 | 0.25 | 1.7 | (Bivins et al., 2017) |
| N fifty value (*Rotavirus*) | Unitless | 0.653 | 8.24 | 58.16 | (Bivins et al., 2017) |
| Ratio *Rotavirus* to E coli | Unitless | 6.055*10^-9^ | 9.533*10^-7^ | 2.5*10^-5^ | (Bivins et al., 2017) |
| Alpha Value (*Campylobacter*) | Unitless | 0.00022 | 0.155 | 6.96 | (Bivins et al., 2017) |
| Nfifty value (*Campylobacter*) | Unitless | 41.30 | 1673 | 35554 | (Bivins et al., 2017) |
| Ratio *Campylobacter* to e coli | Unitless | 6.054*10^-9^ | 6.59*10^-3^ | 1.63 | (Bivins et al., 2017) |

### S2 Figures

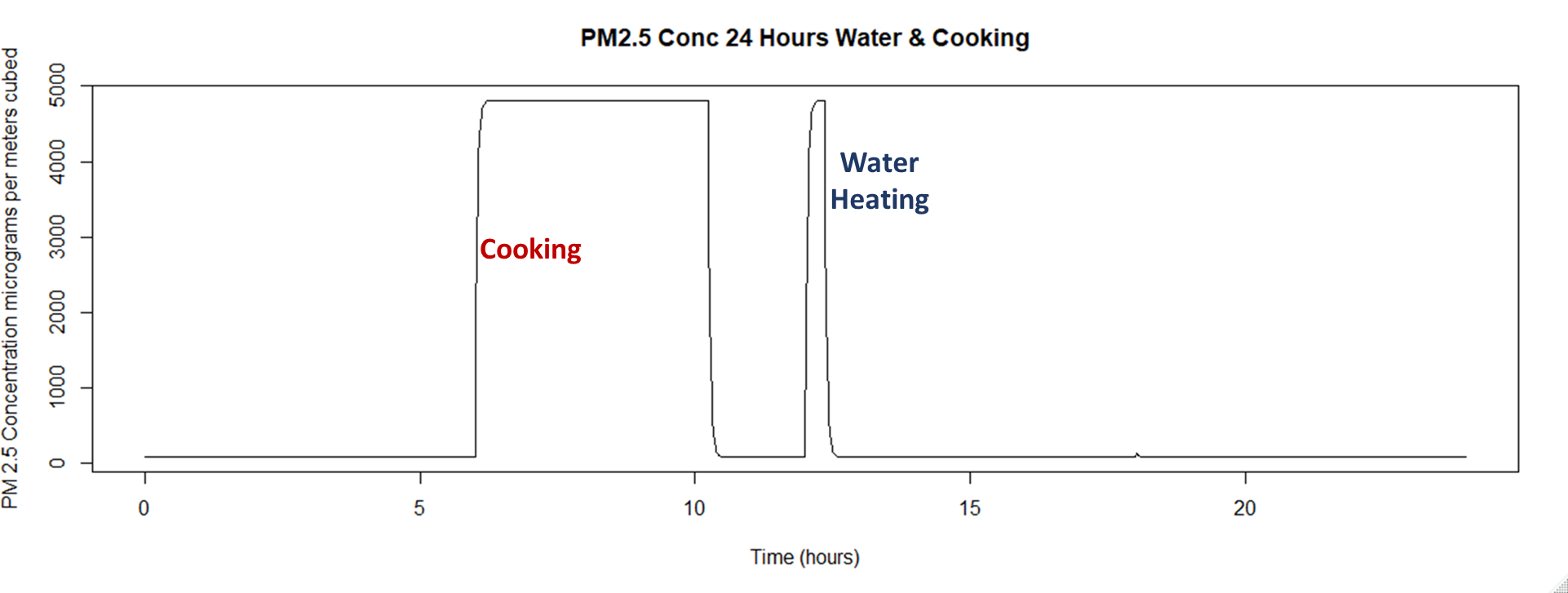

Figure S 1 Example Air Pollution Box Model Run of 24 hour PM_2.5_ Concentration

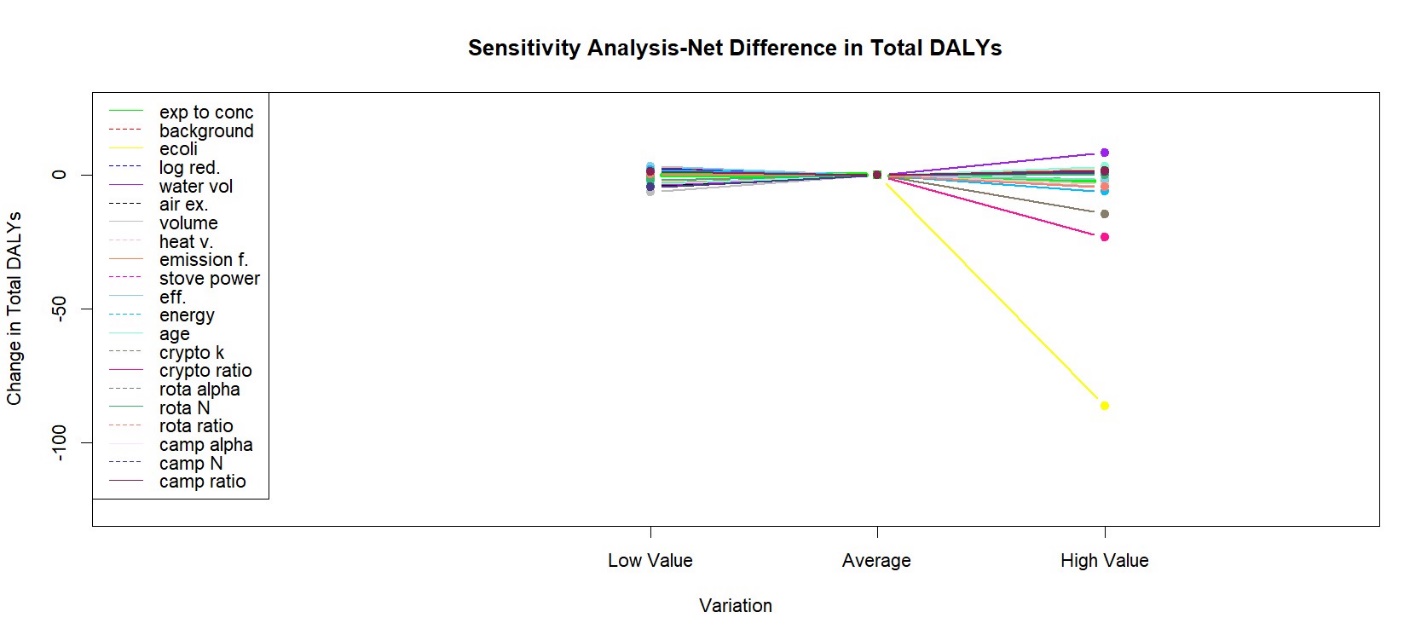

Figure S 2 Sensitivity Analysis Showing the Net Difference in Total DALYs – Total DALYs (defined as the increase in air pollution DALYs from boiling water minus the decrease in DALYs from ) using the lowest, average, and highest values from the parameter distributions, with the average values as a baseline.

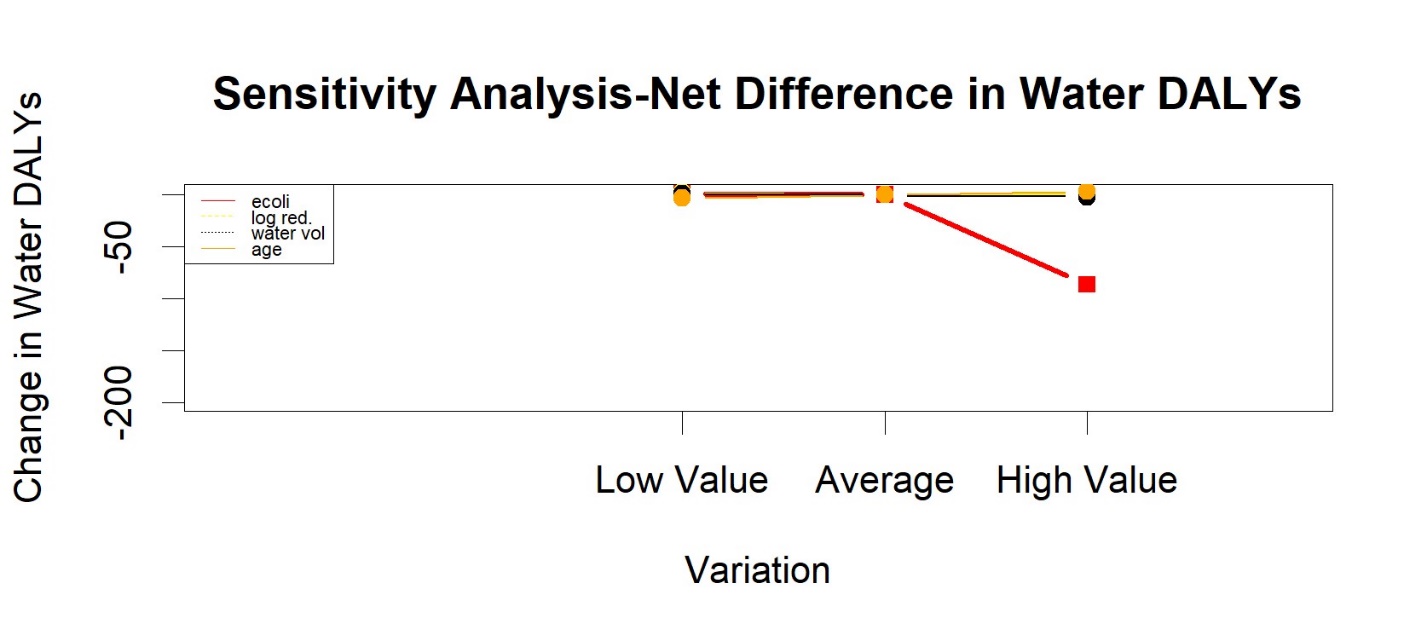

Figure S 3 Sensitivity Analysis For Water DALYs - Showing the Water DALYs (drinking water DALYs after boiling water compared to before boiling) using the lowest, average, and highest values from the parameter distributions, with the average values as a baseline.

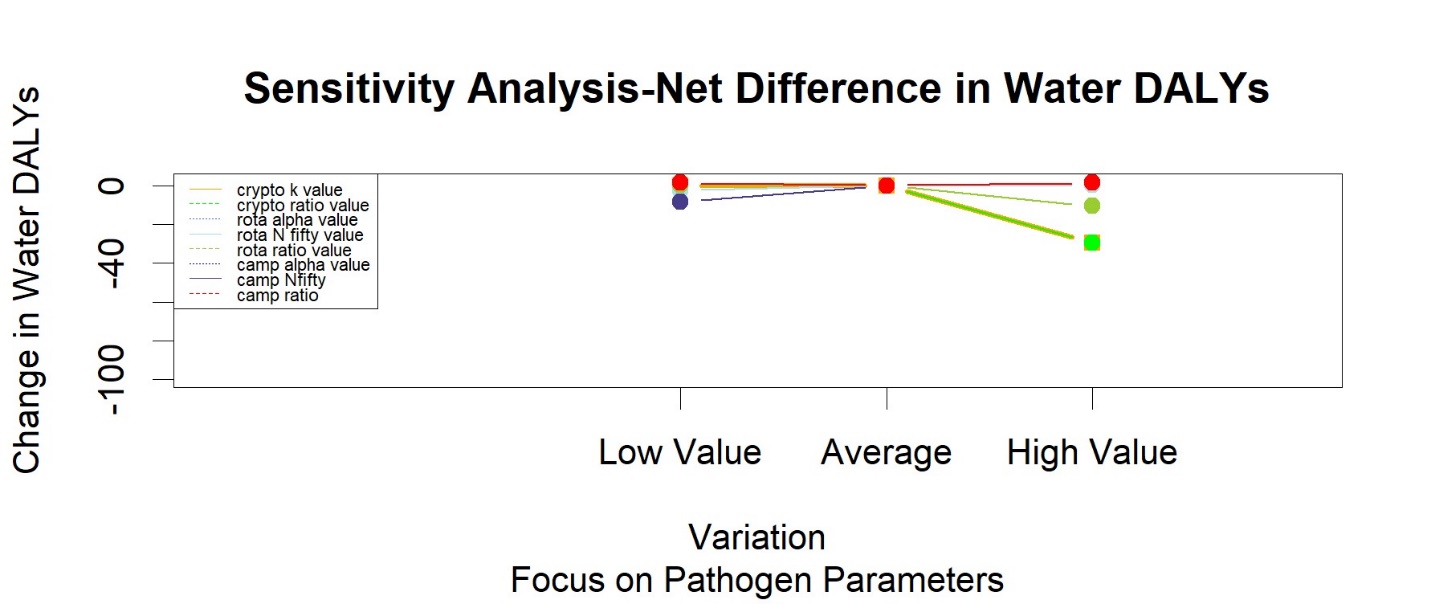

Figure S 4 Sensitivity Analysis for Water DALYs: Pathogen Specific Parameters – Showing the Water DALYs (drinking water DALYs after boiling water compared to before boiling) using the lowest, average, and highest values from the parameter distributions, with the average values as a baseline.

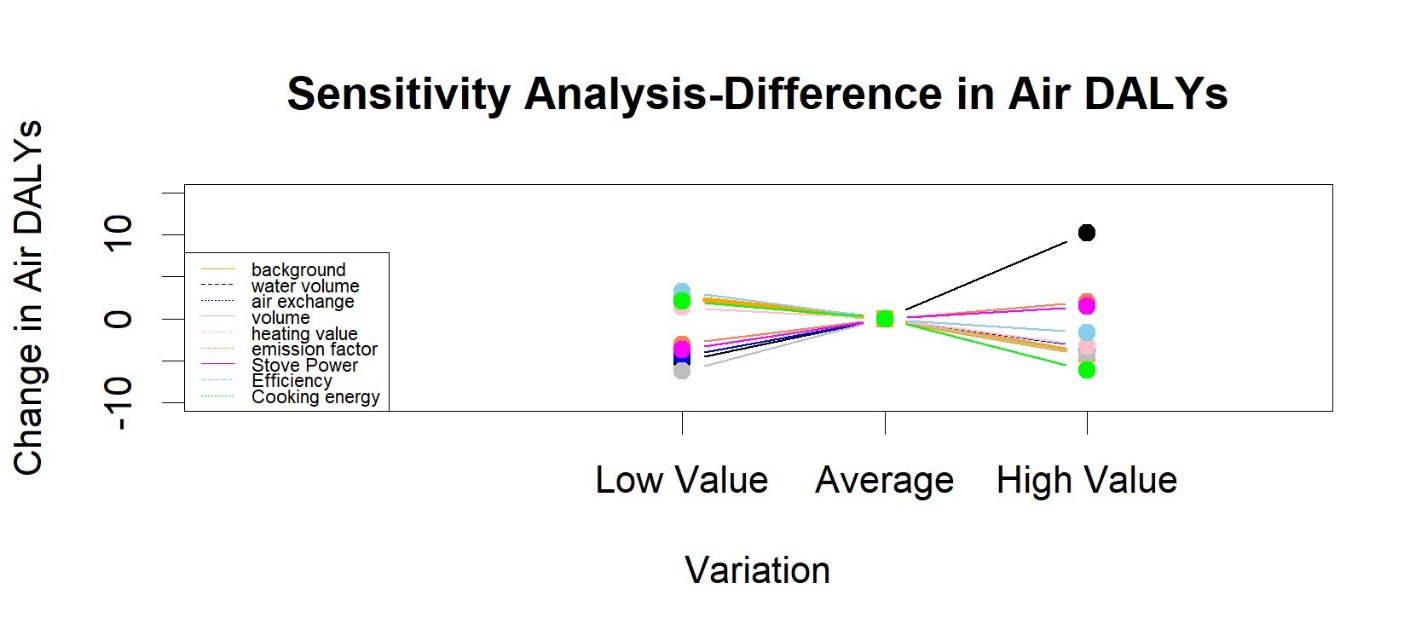

Figure S 5 Sensitivity Analysis Showing in Air DALYs (DALYs from boiling water and cooking compared to just cooking) using the lowest, average, and highest values from the parameter distributions, with the average values as a baseline.
